# Medium and Long Term Time-to-Event Outcomes After Elective Fenestrated and Branched Endovascular Repair of Complex Abdominal and Thoracoabdominal Aortic Aneurysms: A Contemporary Systematic Review

**DOI:** 10.64898/2026.02.08.26345837

**Authors:** Janice Yiu, Mohamed Abdelhalim, Aurelien Gueroult, Imashi Iddawela, Ashish S Patel, Sam Norton, Bijan Modarai

**Affiliations:** Academic Department of Vascular Surgery, South Bank Section, School of Cardiovascular and Metabolic Medicine & Sciences, BHF Centre of Excellence, King’s College London, London SE1 7EH, UK; St George’s Vascular Institute, St George’s, University of London, UK; Department of Inflammation Biology, Department of Psychology, Institute of Psychiatry, Psychology and Neuroscience, Centre for Rheumatic Diseases, King’s College London, UK

**Keywords:** “Aortic Aneurysm, Thoracoabdominal”, “Aortic Aneurysm, Abdominal”, “Endovascular Procedures”, “Survival Analysis”, “Stents” (all MeSH terms)

## Abstract

**Objective:** To define contemporary medium and long term survival and durability outcomes after elective fenestrated and branched endovascular aortic repair (F/BEVAR) for complex abdominal and thoracoabdominal aneurysms and to assess the certainty of the available evidence.

**Data Sources:** MEDLINE, Embase and the Cochrane Library were searched from January 2000 to February 2026, supplemented by citation screening.

**Review Methods:** Published Kaplan Meier time-to-event data were digitised and reconstructed into individual patient datasets. Pooled survival probabilities were generated using validated methods for meta-analytic methods for survival curves. Certainty of evidence was assessed using the GRADE framework.

**Results:** Twenty-four studies comprising 8,886 patients were included. Pooled overall survival was 91.3% (95% CI: 90.7, 91.9) at 1 year, 73.0% (95%CI: 71.9, 74.0) at 3 years and 55.4% (95% CI: 53.9, 56.8) at 5 years. Estimated median overall survival was 6.36 years.

At 5 years, freedom from aneurysm-related mortality was 96.4% (95%CI: 95.3, 97.2), freedom from reintervention was 66.5% (95%CI: 64.6, 68.2), and target vessel patency (TVP) was 94.8% (95%CI: 93.3, 96.0). Certainty of evidence was low for overall survival, aneurysm related mortality and reintervention, and very low for TVP.

**Conclusion:** Elective F/BEVAR provides durable aneurysm exclusion with low aneurysm related mortality; however, long term survival declines substantially. There is a need for more robust survival data and improved tools to support patient selection, shared decision making, and assessment of anticipated benefit when considering prophylactic complex endovascular repair.

**What this paper adds:** This study provides a time-to-event synthesis of medium and long term outcomes after elective F/BEVAR for complex abdominal and thoracoabdominal aortic aneurysms, analysing reconstructed survival data from 8,886 patients across 24 studies published between 2000 and 2025.

This analysis provides empirical survival data to inform recent European Society for Vascular Surgery guidance, demonstrating a median survival of 6.36 years and showing that most late deaths are not aneurysm related. These findings quantify the divergence between procedural durability and long term survival, supporting an individualised treatment strategy grounded in assessment of life expectancy, competing risk and shared decision making.

## Introduction

Evaluating long term outcomes to determine patient benefit from abdominal aortic aneurysm repair is central to contemporary decision making, yet these outcomes remain variable and incompletely characterised^1,2^ This is critical for appropriate selection of patients with complex aortic aneurysms undergoing elective fenestrated and branched endovascular aortic repair (F/BEVAR), which is a technically more demanding intervention than standard repair and is generally associated with higher procedural risk. In the context of prophylactic repair, careful consideration of the balance between procedural risk and anticipated benefit is required^3^.

The European Society for Vascular Surgery (ESVS) 2026 Clinical Practice Guidelines on the Management of Descending Thoracic and Thoraco-Abdominal Aortic Diseases emphasise the need to account for patient longevity, competing risks, and long term outcomes when selecting patients for aortic repair^4^. While this principle is central to practice, there is limited evidence base to support these prognostic judgements. Robust data describing long term survival, late complications, and sustained benefit after aneurysm repair are heterogeneous, and often derived from historical cohorts that may not reflect modern techniques or patient selection. The present evidence synthesis was undertaken to critically evaluate and contextualise the best available time-to-event data from studies published between 2000 and 2025, focusing on elective F/BEVAR for complex abdominal (cAAAs) and thoracoabdominal aortic aneurysms (TAAAs) with medium term (_≤_5 years) and long term (>5 years) follow-up. It aims to provide a clear assessment of current evidence and to define priorities for future research to support more reliable prognostic modelling and personalised decision making in aneurysm care.

## Methods

Search strategy, study selection and data analyses were performed in accordance with the Preferred Reporting Items for Systematic Reviews and Meta-Analyses (PRISMA) guidelines^5^. The protocol was registered prospectively with the International Prospective Register of Systematic Reviews (PROSPERO CRD42024565664). An electronic search of databases (MEDLINE, Embase & Cochrane Library) for human clinical studies published from 2000 to 2026 (updated 7/02/2026) in English was conducted. A combination of medical subject heading (MeSH) terms and keywords were used to maximise sensitivity (full search strategy in Supplementary material). Reference lists for review articles which passed full text review and studies which fit inclusion criteria were searched for relevant studies. Inclusion criteria were studies with published medium to long term outcomes of F/BEVAR of intact TAAAs (Crawford Types I-V)^6^ and cAAAs. These included juxtarenal, pararenal, suprarenal and short-neck infrarenal aneurysms not amenable to standard infrarenal endovascular aortic repair (EVAR)^9^.

Study types included were retrospective observational cohort studies and case series with _≥_50 subjects reporting on elective F/BEVAR for treatment of TAAAs and/or cAAAs. Medium and long term outcomes included as all-cause mortality with a minimum follow-up period of 3 years to 5 years and 5 years or more, respectively. The minimum requirement was for a study to include Kaplan-Meier survival analysis or a cumulative incidence function (CIF) for all-cause mortality. The primary outcome was all-cause mortality but expressed as overall survival for the analysis. Secondary outcomes were freedom from aortic related mortality, freedom from reintervention and target vessel patency. Major reinterventions were defined as deployment of proximal or distal extensions with large diameter (>12F) sheaths, removal of stent graft, use of thrombectomy or thrombolysis in target vessels, and any open surgical procedure to treat underlying aortic disease^7^.

Exclusion criteria were animal studies, conference abstracts, case reports and case series with fewer than 50 patients and aneurysm pathologies that were non-degenerative, such as mycotic or post dissection aneurysms. Studies where >5% of patients underwent urgent, or emergency aneurysm repairs were excluded. For publications of the same population from the same centre, the publication with longest follow-up duration was included.

J.Y., M.A., I.I. screened abstracts and selected studies independently for inclusion with the Rayyan tool^8^. Any discrepancies in judgement regarding search strategy, paper selection, quality assessment and data extraction were resolved through discussion between all authors.

Study quality and risk of bias assessment were conducted using the ROBINS-I tool (version 2)^9^ by J.Y. and M.A. The certainty of results was rated using the Grading of Recommendations Assessment, Development and Evaluation (GRADE) approach (J.Y. and M.A.), which provides a structured framework for rating confidence in effect estimates across studies. GRADE is used to classify the overall certainty of evidence as high, moderate, low, or very low by evaluation of study design, risk of bias, inconsistency, imprecision and publication bias^10^.

Using a standardised data extraction form, J.Y. and M.A. extracted data from included studies. Data extracted from the included studies were publication year, study design, number of patients, demographics, anatomical aneurysm classification, maximum aortic diameter, stent graft type (i.e. off the shelf or custom-made, fenestrated or branched), number of target vessels, intraoperative data and reinterventions.

For Kaplan-Meier curves of time-to-event data, raw patient data were extracted from these curves using the open source digitize software^11^. Individual patient data (IPD) were generated from data points including numbers at risk, risk time (years), length of time interval (for reporting survival probability) and vector of survival probabilities in open source IPDfromKM software^12^.

Categorical variables were expressed as counts and percentages, and continuous variables as mean with standard deviation. Median and interquartile range values were converted to approximate mean and standard deviations using the method described by Wan et al^13^. Meta-analyses of Kaplan-Meier estimated probabilities was undertaken, applying methodology described by Combescure et al^14^. Summary Kaplan-Meier curves for overall survival, freedom from aneurysm related mortality, freedom from reintervention and target vessel patency were plotted from pooled probabilities in STATA 18 software (StataCorp, Texas, 2025)^15^. Data is mature up to the point where only 10% of numbers at risk at T_0_ remains in the data analysis.

Mixed-effects parametric survival models were fitted using STATA’s mestreg command, with shared frailties for each study. The functional form for survival function included consideration of Weibull, exponential, log-logistic, lognormal and gamma. The best fitting model was selected based on the Bayesian Information Criterion (BIC), with the model exhibiting the lowest BIC considered optimal for describing the survival data. To assess between-study heterogeneity in time-to-event outcomes, a mixed-effects Weibull survival model was fitted, with random intercepts for each study to account for clustering and unobserved heterogeneity. Anatomical aneurysm classification was included as a dummy coded fixed effect. Model fit was assessed using a likelihood ratio test comparing the mixed-effects model with a fixed-effects Weibull model. Between-study heterogeneity was considered statistically significant at *p* < .05.

For studies which published Kaplan-Meier curves in separate groups instead of an overall single curve, e.g. patients grouped into males and females, grouped into under 80 years and over 80 years, raw patient data and IPD were generated from these individual curves and pooled together as one dataset per study for analysis. For studies that published CIF instead of Kaplan-Meier time-to-event curves, the digitization and IPDfromKM were not used due to risk of overestimation of event-free survival. Survival probabilities were approximated from the Fine & Grey approach as 1-KM survival (i.e., survival ∼ 1 – CIF)^16^. This approach assumes that the rate of competing events was low and that the cumulative incidence closely approximates event-free survival.

Due to the variability in aneurysm anatomies reported in the studies, outcomes were analysed as an overall cohort and two separate subgroups determined by anatomical aneurysm classification: (1) studies including both cAAAs and types I-V TAAAs; (2) studies including cAAAs only; (3) studies including types I-V TAAAs only.

## Results

A total of 1330 studies were generated from initial electronic search, in which 930 studies were screened after removal of duplicates. Including citation search, a total of 141 studies were chosen for full text screening with 24 studies eventually included for meta-analysis. The quality and risk of bias of included studies were assessed with Cochrane ROBINS-I (V2) tool^9^ and presented in Supplementary Material.

### Demographics

A total of 24 studies comprising 8886 patients who underwent F/BEVAR were included in the analysis. The pooled mean age was 73.9±7.7 years and 6593 (74.7%, 95% confidence interval [CI]: 73.8-75.6) were males. Out of 4453 patients with cAAAs, there were 78.6% (3502) juxtarenal aneurysms, 8.5% (377) short-neck infrarenal aneurysms, 7.7% (341) pararenal and 5.2% (233) suprarenal aneurysms. There were 4433 patients reported with TAAAs, with individual numbers not provided in two studies for Types I-III^18^ and Types I-V^19^. For studies that reported individual numbers, there were 5.9% (182) Type I, 19.8% (615) Type II, 13.8% (429) Type III, 59.1% (1832) Type IV and 1.4% (43) Type V TAAAs. The mean follow-up time was 4.3 ± 1.6 years.

With the ROBINS-I(v2) tool, 17 out of 24 studies were judged to have low overall risk of bias and 7 as moderate risk. Demographics, comorbidities, and early post-operative outcomes of individual studies are presented in Supplementary Material.

## Overall survival

### All included studies

There were 24 studies with 8886 patients in total with six long term studies (more than 5 years)^20–25^, and eighteen medium term studies (two studies with 5 years follow-up^26–27^, sixteen studies with 3 years but less than 5 years follow-up)^28–42^. The pooled Kaplan–Meier survival estimates demonstrated an overall survival probability of 91.3% (95%CI: 90.7, 91.9) at 1 year, 73.0% (95% CI: 71.9, 74.0) at 3 years and 55.4% (95% CI: 53.9, 56.8) at 5 years. With a 10% threshold, data maturity reached 5 years (Figure 2). GRADE certainty of evidence was low. Based on the Weibull parametric survival model, the overall estimated median survival time for all patients included in this meta-analysis is 6.36 years (95%CI: 5.48, 7.25).

**Figure 1.**
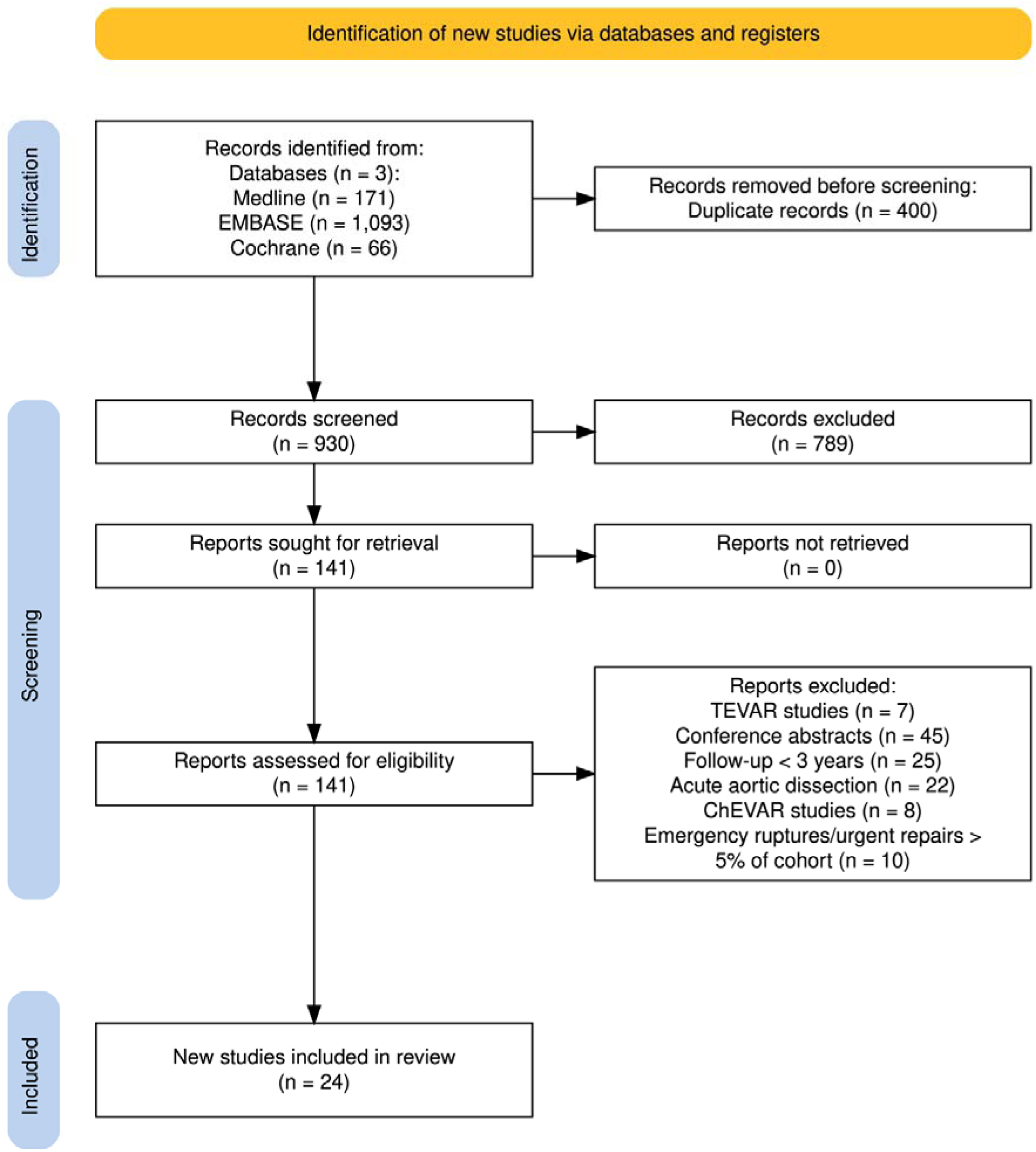
The Preferred Reporting Items for Systematic reviews and Meta-analyses (PRISMA) flow diagram showing the search, screening and selection process for included studies of F/BEVAR of intact thoracoabdominal and/or complex abdominal aortic aneurysms^17^.

**Figure 2.**
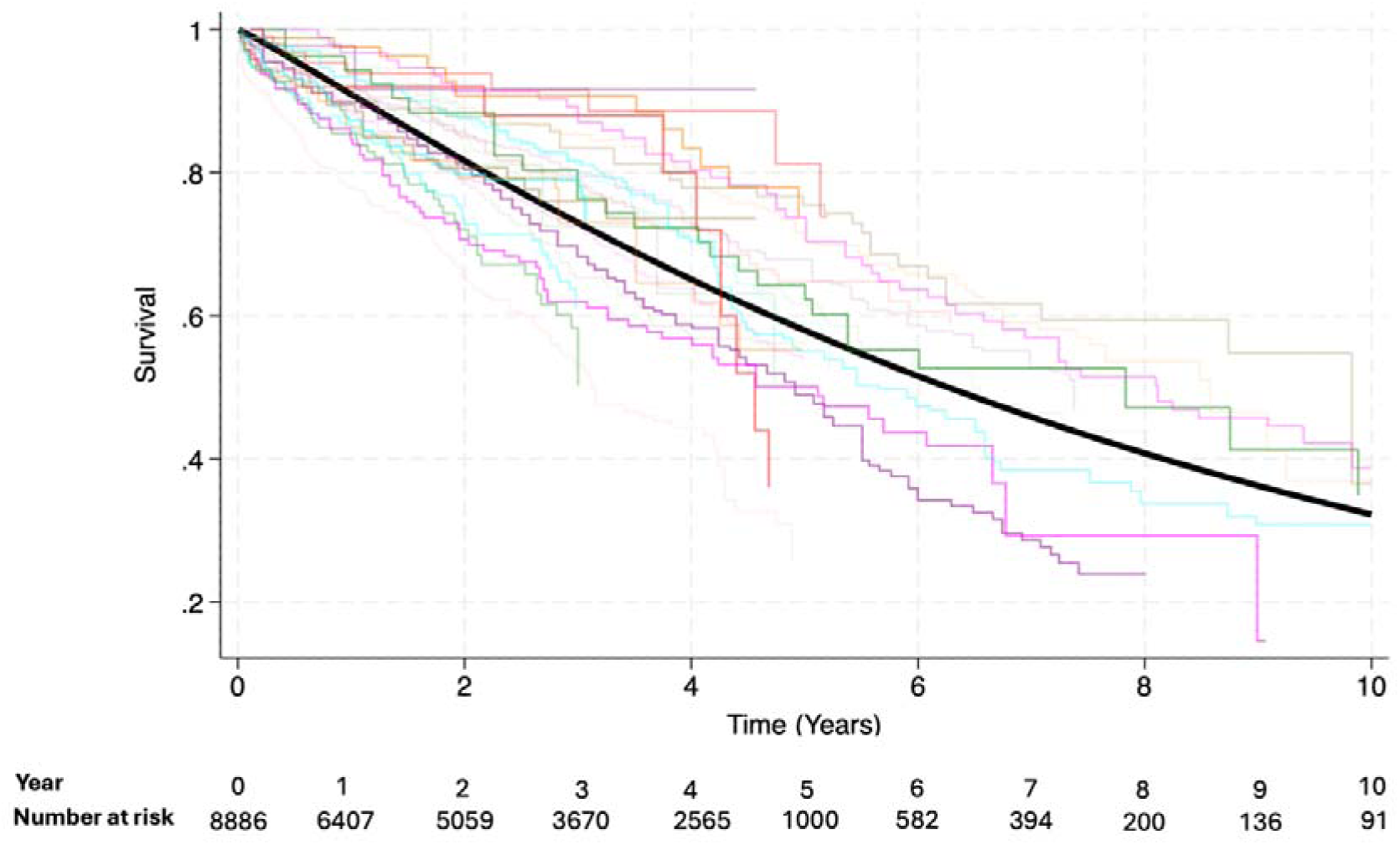
Kaplan-Meier overall survival estimates of all included studies The pooled Kaplan-Meier survival estimate is shown in black with individual Kaplan-Meier survival estimates of all included studies presented ^20–42^. Key for coloured curves of individual studies omitted due to large number of studies for presentation.

### Complex abdominal aortic aneurysms

From a total of 1403 patients from 8 studies^20,23,26,31,34,37,40,42^, the pooled Kaplan-Meier survival estimates after F/BEVAR of cAAAs were 93.6% (95%CI: 91.3, 94.9) at 1 year, 78.7% (95% CI: 76.2, 81.0) at 3 years, 61.5% (95% CI: 58.4, 64.5) at 5 years and 42.5% (95%CI: 38.5, 46.5) at 8 years (Figure 3) and maturity of data reached 8 years. From the Weibull estimated survival model, the overall median survival was 7.22 years (95% CI: 5.80, 8.64).

**Figure 3.**
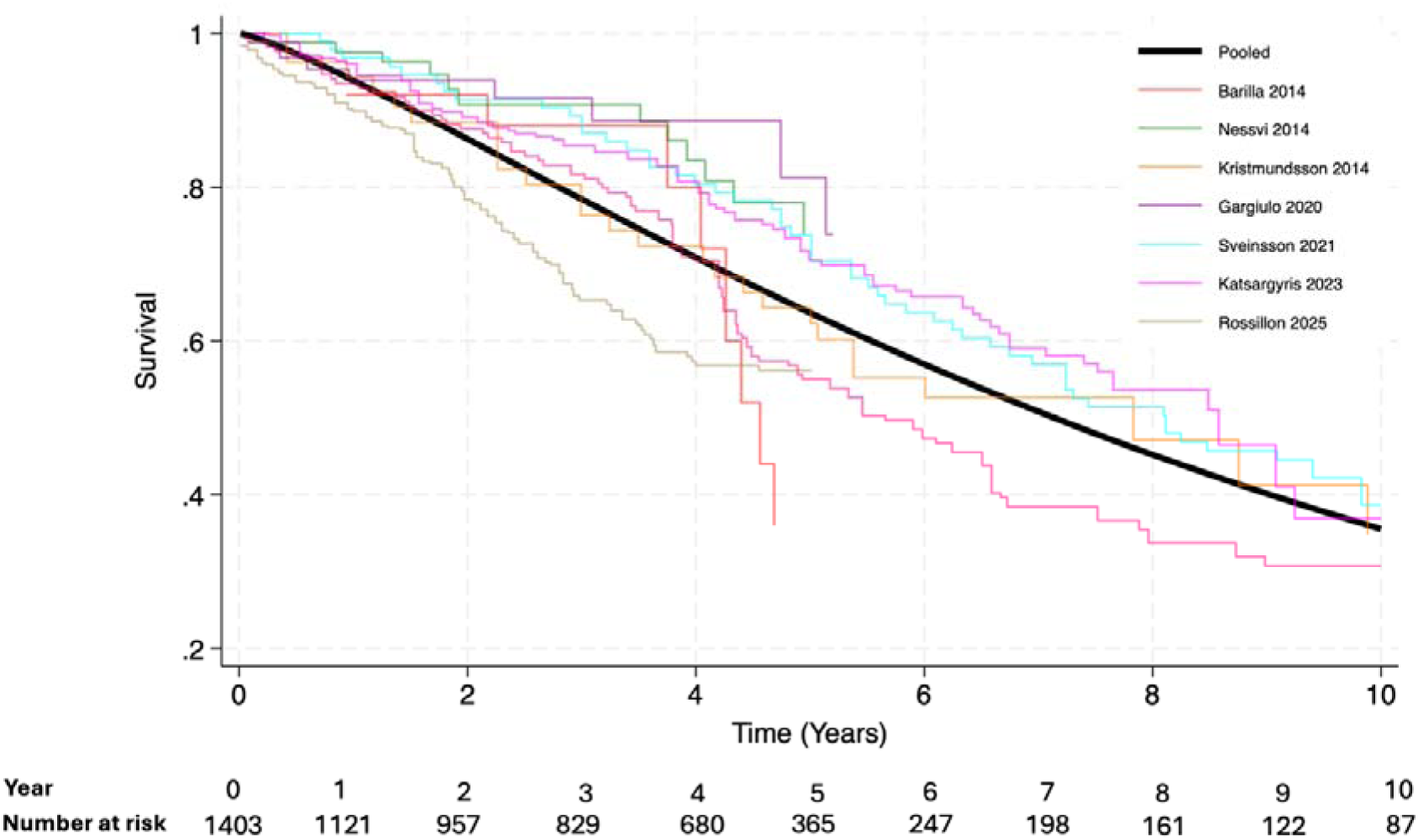
Kaplan-Meier overall survival estimates of studies that included F/BEVAR of complex abdominal aortic aneurysms (cAAAs) only The pooled Kaplan-Meier survival estimate is shown in black with individual Kaplan-Meier survival estimates of studies that only reported on cAAAs presented^20,23,26,31,34,37,40,42^.

### Thoracoabdominal aneurysms

From 2459 patients in 3 studies, reporting on Types II & III^30^, Types I-III^18^ and Types I-IV^19^ TAAAs, the pooled Kaplan-Meier survival estimates were 88.6% (95%CI: 87.3, 89.8) at 1 year and 73.0% (95%CI: 71.0, 74.8) at 3 years (Figure 4). The Kaplan-Meier curves were truncated at the last time point with patients at risk to avoid extrapolation beyond observed data, as number at risk fell to zero beyond time point of 4 years.

**Figure 4.**
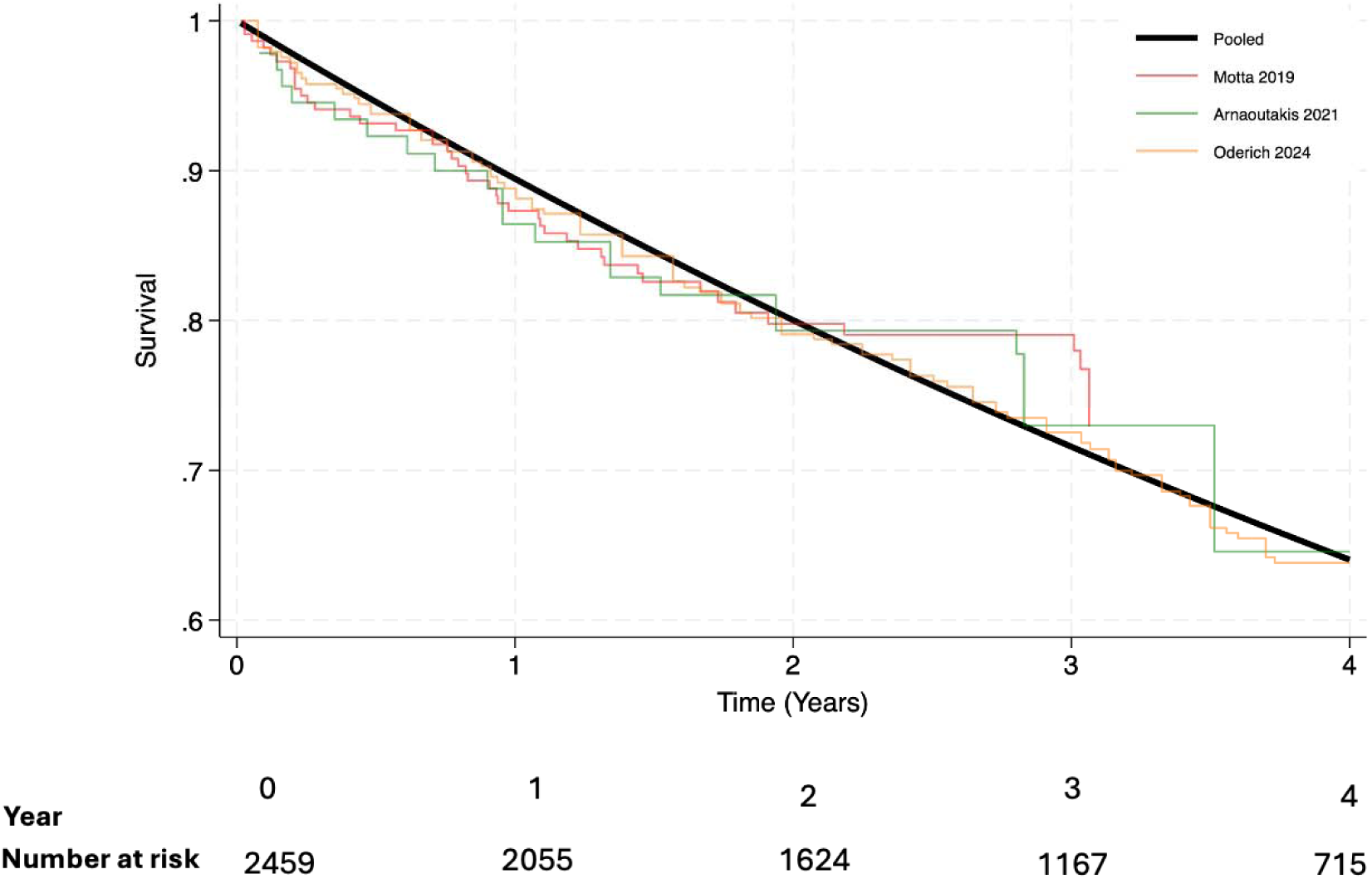
Kaplan-Meier overall survival estimates of studies reporting on F/BEVAR of thoracoabdominal aneurysms (TAAAs) only The pooled Kaplan-Meier survival estimate is shown in black with individual Kaplan-Meier survival estimates of studies that only reported on TAAAs^18,19,30^.

### Between-study heterogeneity

A mixed-effects Weibull proportional hazards model was used to compare overall survival across aneurysm subtypes (mixed aneurysm group, TAAAs only, cAAAs only), adjusting for between-study heterogeneity via random intercepts for study ID. The log-likelihood ratio (LR) test comparing the mixed-effects model to a standard Weibull model was significant (chibar² (01) = 214.23, *p < .001*), indicating substantial between-study heterogeneity, and supporting the inclusion of random effects. The estimated variance of the random intercept was 0.119 (95% CI: 0.06, 0.25), reflecting moderate heterogeneity across studies.

Compared with the reference group (TAAAs only group), the hazard ratio (HR) for overall survival in the cAAAs only group was 0.75 (95%CI: 0.45, 1.24; *p = .26*), and in the mixed aneurysm group was 1.03 (95%CI: 0.64, 1.66, *p = .90*). These differences were not statistically significant, suggesting no strong evidence of variation in survival across the three aneurysm subtypes after adjusting for study-level clustering.

The baseline hazard parameter was significant (HR = 0.096, 95%CI: 0.06, 0.15; *p < .001*), and the shape parameter of the Weibull model was positive (ln(p) = 0.102), indicating an increasing hazard over time.

### Freedom from aneurysm-related mortality

Freedom from aneurysm-related mortality was reported in a Kaplan-Meier graph in only four studies^24,29,37,38^and cumulated incidence function (CIF) in two studies^39,41^. One study reported on TAAAs^41^, one reporting on cAAAs^37^ and four studies reported on cohort of mixed types ^24,29,38,39^. The pooled estimates are based on these studies, with a combined total of 2818 patients.

The pooled Kaplan–Meier survival estimates demonstrated an overall freedom from aneurysm related mortality of 98.8% (95%CI: 98.3, 99.2) at 1 year, 98.2% (95%CI: 97.6, 98.7) at 3 years and 96.4% (95%CI: 95.3, 97.2) at 5 years post intervention (Figure 5) and data maturity was reached at 6 years. The GRADE certainty of evidence was rated as low.

**Figure 5.**
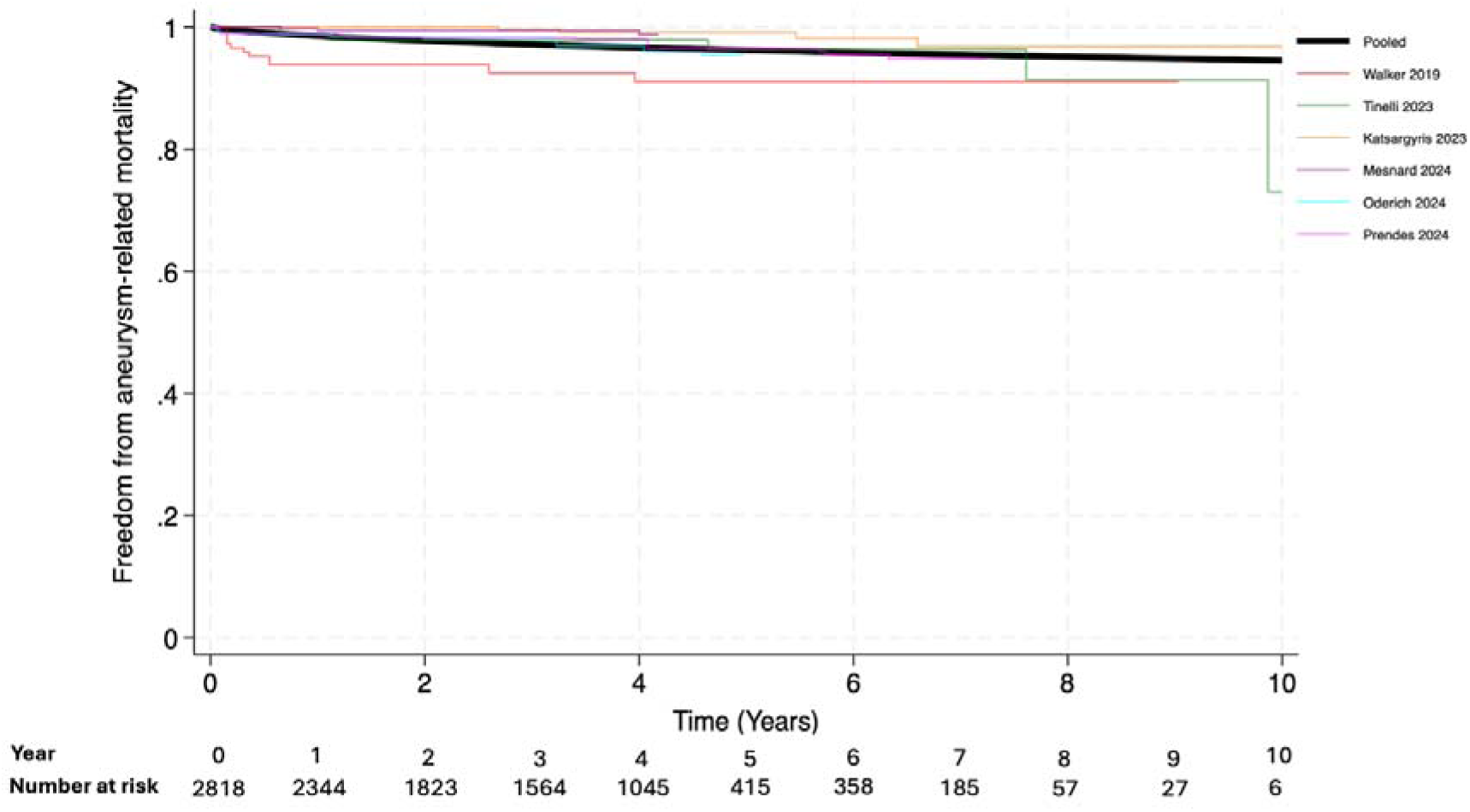
Pooled versus individual freedom from aneurysm-related mortality Kaplan-Meier curves of studies reporting on F/BEVAR of complex abdominal and thoracoabdominal aneurysms (cAAAs and TAAAs) The pooled freedom from aneurysm-related mortality is shown in black with individual freedom from aneurysm-related mortality estimates of one study that reported on cAAAs only^37^, one study that reported on TAAAs only^41^ and four studies that reported on combined cohort of cAAAs and TAAAs^24,29,38,39^.

Parametric modelling was tested, but overestimated survival, showing similar miscalibration once extrapolated beyond maximum median follow-up of 10 years. Consequently, model derived median survival time was not presented for this outcome.

### Freedom from reintervention

Freedom from reintervention, presented as Kaplan-Meier analyses, was reported in fourteen studies and cumulative incidence function in one study^41^ with a total of 4555 patients included. Of these, five studies reported on cAAAs only^20,23,32,37,40,42^, two studies reported on TAAAs only^19,41^and seven reported on mixed cohort of complex abdominal aortic and thoracoabdominal aneurysms^21,24,27,32,33,36,38^.

The pooled freedom from reintervention was 85.4% (95%CI: 84.4, 86.5) at 1 year, 73.6% (95%CI: 72.1, 75.0) at 3 years and 66.5% (95%CI: 64.6, 68.2) at 5 years when data maturity has been reached (Figure 6). The GRADE certainty of evidence was rated as low.

**Figure 6.**
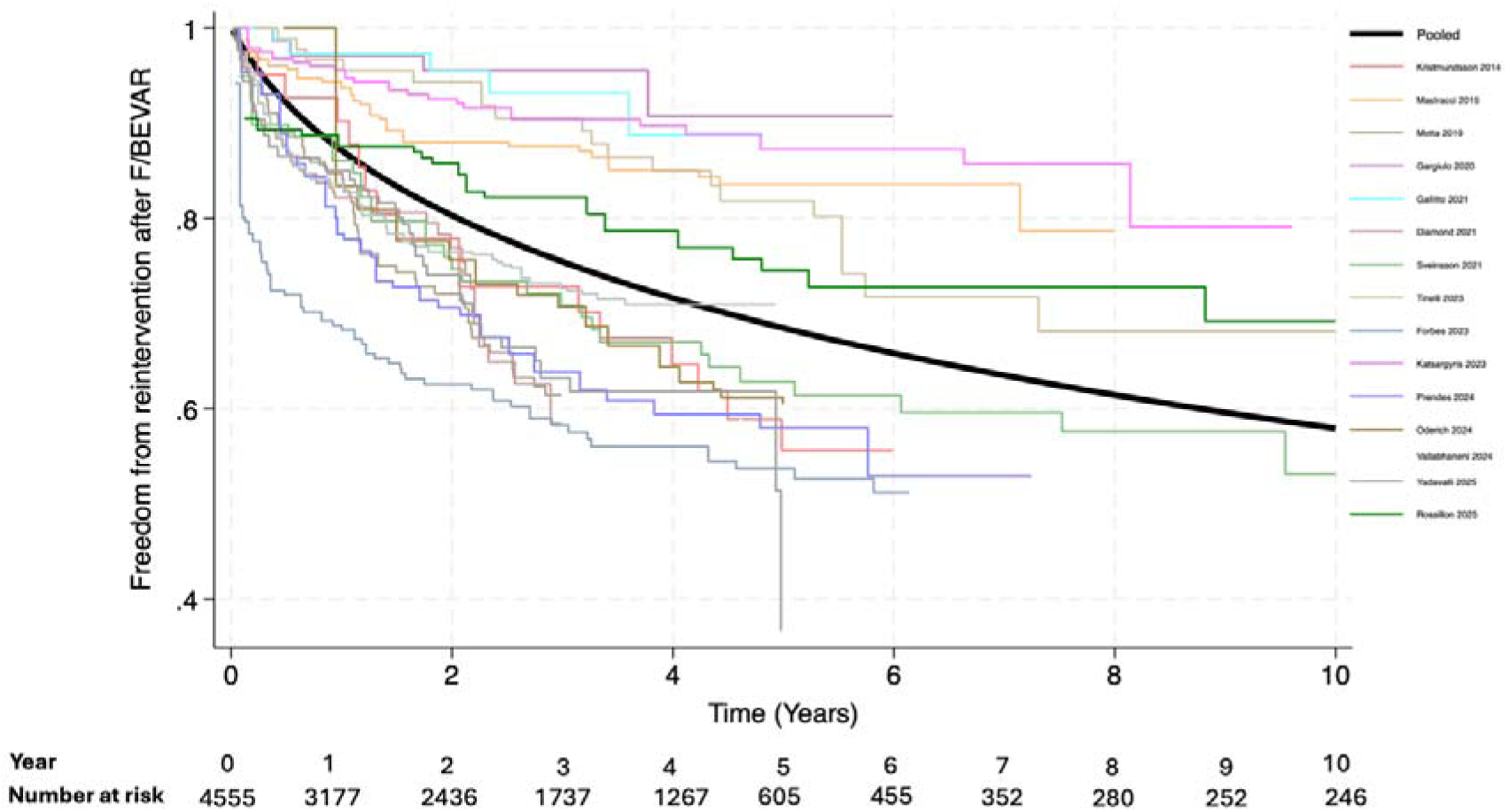
Pooled versus individual freedom from reintervention Kaplan-Meier curves after F/BEVAR of complex abdominal aortic and thoracoabdominal aneurysms (cAAAs and TAAAs) The pooled freedom from reintervention after F/BEVAR repair of cAAAs and TAAAs is shown in black with individual Kaplan-Meier curves shown^19,29,21,23,24,27,31,32,33, 37,38,40,41,42^.

For studies that reported on cAAAs only^20,23,32,31,37,40,42^, the pooled freedom from reintervention Kaplan-Meier survival was 89.1% (95%CI: 87.0, 90.9) at 1 year, 80.4% (95%CI: 77.7, 83.7) at 3 years, 75.4% (95%CI: 72.4, 78.2) at 5 years and 69.0% (95%CI: 64.4, 72.9) at 10 years (Supplementary material). For studies that reported on TAAAs only^19,41^, the pooled freedom from reintervention Kaplan-Meier survival was 83.5% (95%CI: 81.2, 85.6) at 1 year and 63.0% (95%CI: 60.0, 66.0) at 3 years (Supplementary material).

Overall, 14% (845/5900) of patients across eighteen studies underwent reinterventions after F/BEVAR. Six studies provided no absolute number for reinterventions ^22,28,30,35^ or no number of patients who underwent reinterventions^21^. Among reported indications for reintervention, type II endoleak demonstrated highest crude pooled incidence (8.6%), followed by Type III endoleak (7.0%) and graft limb stenosis/occlusion (6.9%).

13% (175/1359) of patients underwent reinterventions from studies that reported exclusively on cAAAs ^20,23,26, 31,37,40,42^with majority of reinterventions for graft limb stenosis/occlusion (7.9%). For studies that reported exclusively on TAAAs^19,41^, 11% patients required reinterventions (143/1285), with insufficient data to comment on frequency of types of reinterventions. For studies that reported outcomes for both cAAAs and TAAAs (including types I-V) ^24,25,27,29,32,33,34,36,38,39^, 16% (530/3256) patients required reinterventions, with a majority for type II (12.2%) followed by type III endoleak (10.7%). The limited number of studies reporting outcomes in cAAAs only and TAAAs only cohorts precluded formal assessment of between-study heterogeneity.

### Target vessel patency

Complete Kaplan-Meier analyses of target vessel patency were reported in 2 studies^23, 37^, both on cAAAs only cohorts with a total of 1625 vessels. The pooled target vessel patency was 97.6% (95%CI: 96.7, 98.3) at 1 year, 95.4% (95%CI: 94.0, 96.4) at 3 years, 94.8% (95%CI: 93.3, 96.0) at 5 years and 92.6% (95%CI: 90.4, 94.3) at 9 years when data maturity has been reached (Supplementary material). The GRADE certainty of evidence was rated as very low.

### Reported causes of death

Discrete data on late causes of death were reported in 5 studies^31,36,38,39,41^. The most frequent was unknown (64.1%), followed by cardiovascular (22.5%), malignancy (8.4%), pulmonary (2.8%), and cerebrovascular causes (2.2%).

## Discussion

This meta-analysis of time-to-event data provides insights into medium and long term outcomes following elective F/BEVAR of cAAAs and TAAAs. The aggregated results from >8,000 patients demonstrate that early postoperative survival is excellent, but long term survival is progressively diminished. Overall survival probability of approximately 91% at 1 year, declining to 55% at 5 years after F/BEVAR, albeit informed by limited data and maturity. In practical terms, more than half of patients treated with F/BEVAR for cAAAs and TAAAs have died by the 5 years, despite the aneurysm being successfully excluded. This underscores that patient comorbidities and advanced age exert a major impact on long term mortality in elective F/BEVAR^43^. The estimated median survival of 6.3 years after F/BEVAR highlights that many patients undergoing these technically advanced repairs have an inherently limited life expectancy, a crucial consideration when recommending treatment. Only two studies reported on medications aimed at modification of cardiovascular risk factors ^27,28^. The recent recommendations in the European Society for Vascular Surgery (ESVS) 2026 suggest that addressing these risk factors should become an area of greater focus^4^.

Despite the need for reinterventions, it is encouraging that aneurysm exclusion appears durable in most patients, as evidenced by the relatively low incidence of aneurysm-related mortality over time, with a 96% freedom from aneurysm-related mortality at 5 years. Our analysis was limited in determining the exact causes of late mortality, with granular cause of death data reported^31,36,38,39,41^. In those that did, deaths due to cardiovascular causes and malignancies were prominent, and a large proportion remained unspecified. The high prevalence of “unknown” causes of death reflects real-world challenges in long term follow-up of vascular patients, especially when deaths occur outside the hospital. This finding echoes concerns raised by Whittaker et al^44^ about difficulties obtaining cause of death information for patients after abdominal aortic aneurysm interventions and lack of postmortem examination and thus, definitive evidence of causes of death^45^. More diligent tracking of late mortality, for example, via national death registries, would help clinicians understand what ultimately claims patients’ lives after elective aneurysm repair.

From a decision making standpoint, F/BEVAR extends the option of aneurysm repair to patients who might be poor candidates for open surgery, offering a chance to prevent rupture with an endovascular approach. However, the data suggest that many of these patients will not survive beyond 6 years, due to other comorbidities. Current guidelines emphasise considering patient life expectancy in offering AAA repairs. The 2026 ESVS guidelines^4^, with a new Class IIIB recommendation, do not advise elective endovascular surgical repair for TAAAs in patients with ‘*limited life expectancy*’. Our analysis affirms that guidance: in this cohort, the median survival of approximately 6 years, is well above that threshold, indicating that, on average, patients selected for F/BEVAR had a life expectancy justifying intervention. However, the wide range of outcomes (with a quarter of patients dying by ∼3 years after F/BEVAR) means that some likely underwent a complex, costly intervention with very few life years gained. Identifying those patients preoperatively is challenging. Comprehensive geriatric and cardiovascular risk assessment should therefore be part of the workup for F/BEVAR^45^. Tools to gauge frailty and end organ function can help prognosticate whether a given patient is likely to enjoy the benefit of aneurysm repair for several years, or conversely, whether they are so high risk that a conservative approach might be more prudent. Emerging strategies like prediction models and machine learning applied to large vascular datasets may aid risk stratification. For example, a recent machine learning analysis of over 10,000 patients in the Vascular Quality Initiative (VQI) registry showed promise in predicting 1-year outcomes after cAAAs repairs^46^. While such models are in early stages, they could eventually complement clinical judgment in tailoring patient selection.

Another consideration is the resource commitment that F/BEVAR entails. Evidence on the cost effectiveness of elective F/BEVAR remains limited. A scoping review of seven observational and registry-based studies on intact cAAAs (2006–2014) reported that the principal driver of higher costs compared with open repair was the price of stent grafts^47^. Additionally, our findings show that reintervention after F/BEVAR is not uncommon. For studies that reported sufficient detail on reinterventions, 1 in 8 patients with cAAAs required reintervention, compared to 1 in 9 patients with TAAAs and 1 in 6 for studies that analysed both types of aneurysms as a cohort. This highlighted the importance for future studies to present findings for cAAAs and TAAAs separately for more accurate evaluation of reintervention rate.

Although most reinterventions were endovascular and technically manageable, evidence suggested that reinterventions do not impact long term survival, but could impose additional burden^48,49^. Prior analyses have questioned whether F/BEVAR is justified in patients with very limited longevity, given the high upfront device cost, intensive postoperative follow-up and potential need for reintervention^47^. A meta-analysis of time-to-event data of patients treated electively with custom-made FEVAR for cAAAs and types IV TAAAs from studies published up to 2023 noted insufficient data on long term outcomes beyond 5 years^50^. Our study suggested that for patients with a reasonable life expectancy, F/BEVAR offers a chance of long term aneurysm related survival (about 96% at 5 years freedom from aneurysm related death) and relatively low risk of rupture if surveillance is maintained.

Some important outcomes remain underreported in the literature and thus in our meta-analysis, including the quality of life (QoL) after F/BEVAR. Only one study^25^ systematically assessed QoL at 5 years (using SF-36), leaving a gap in understanding the functional status and life quality of survivors. As more patients undergo complex endovascular repairs, incorporating patient reported outcomes and QoL measures into studies will be valuable^51^. Similarly, outcomes pertaining to radiation exposure and risk of malignant transformation, remain poorly reported. This is notable given that cancer was the third most common cause of death in the studies analysed, and a previous study has suggested higher cancer-related mortality after EVAR compared with open repair^52^. There remains insufficient consistency in reporting cumulative radiation doses, including resultant exposure from follow-up surveillance CT scanning. Using standardised dose reporting to potentially correlate radiation exposure with late adverse events in the future is an area of unmet need.

## Limitations

All included data were from non-randomised series, predominantly retrospective cohort studies. Consequently, the certainty for all outcomes was rated low to very low. Substantial number of clinical studies were excluded due to lack of follow-up data beyond 2 years including a few from the US Aortic Research Consortium^53,54^. Between study heterogeneity was substantial, driven by differences in aneurysm extent and type and reporting practices with several studies that presented findings for mixed cohort of cAAAs and different types of TAAAs. This restricted the interpretability of aneurysm specific subgroup analyses and was mitigated by pooling Kaplan-Meier estimates of secondary outcomes for studies that reported on a specific type of aneurysm. Long term follow-up was limited and studies that did likely represent higher volume centres with better outcomes, which may have biased estimates towards more favourable outcomes. There was a lack of access to individual patient data from original studies. This prevented adjustment for patient level factors such as age, comorbidities and aneurysm type. Any comparisons between subgroups must be interpreted cautiously, as they are unadjusted and prone to bias. Studies that reported secondary outcomes only as point estimates at fixed time intervals, rather than Kaplan-Meier or cumulative incidence function curves could not be included in the pooled time-to-event analysis. Meta-analysis of proportions was not performed, as this approach does not account for censoring or variable follow-up durations, leading to biased and non-comparable pooled estimates. Lastly, there was inconsistency in outcome definitions among studies. A few reported CIF of mortality (accounting for competing risks). While this introduces inconsistency, a uniform analytical approximation was applied to maintain coherence across the data analysis. The assumption that the rate of competing events was low, and that CIF closely approximated event free survival, may not always hold, as it assumes individuals experience only a single cause specific event with the rate of competing risk of mortality being high in this population. While this approximation is generally acceptable when competing events are infrequent, this should be considered when interpreting the results.

## Conclusion

This synthesis of available evidence indicates that procedural success is maintained in the medium term, with high target vessel patency and acceptable reintervention rates, but survival progressively declines due to underlying risk factors and comorbidities. F/BEVAR should be offered to those who stand to benefit by surviving long enough and judiciously avoided in those with curtailed life expectancy. Diligent lifelong surveillance is essential to manage endograft related complications. The vascular community would benefit from standardised reporting of outcomes in rigorously maintained prospective registries stratified by aneurysm type and patient risk factors, as well as innovative approaches, including big data and artificial intelligence to refine prognostication and inform precision decision making.

## Data Availability

All data produced in the present study are available upon reasonable request to the authors.

