## Supplementary material for "Medium and Long Term Time-to-Event Outcomes After Elective Fenestrated and Branched Endovascular Repair of Complex Abdominal and Thoracoabdominal Aortic Aneurysms: A Contemporary Systematic Review"

- Search strategy
- Demographics table
- Table of co-morbidities, intra-operative characteristics, post-operative complications and definition of reinterventions
- Robins-I(v2) table
- GRADE analysis table
- Figure (1): Pooled versus individual Kaplan-Meier estimate of freedom from reintervention after F/BEVAR of complex abdominal aortic aneurysms.
- Figure (2): Pooled versus individual Kaplan-Meier estimate of freedom from reintervention after F/BEVAR of thoracoabdominal aneurysms

Search strategy:

Embase

1. Exp abdominal aorta aneurysm/ or exp abdominal aortic aneurysm/
2. Limit 1 to (human and English language)
3. Exp endovascular surgery/
4. Limit 3 to (human and English language)

Ovid Medline

1. Exp. Aortic Aneurysm, Abdominal
2. Exp. Endovascular procedures
3. Limit 1 and 2 to English Language and Humans
4. Exp survival analysis/ or exp disease free survival/ or exp survival/ or exp mean survival time/ or exp overall survival/ or exp median survival time/ or exp disease specific survival/ or exp event free survival/ or exp progression free survival/ or exp long term survival/
5. Mortality/ or “cause of death”/ or fatal outcome/ or survival rate/
6. Limit 4 and 5 to English language and Humans
7. 3 AND 6

Cochrane Central registry

1. Cochrane Central Register of Controlled Trials

All text

Aortic abdominal aneurysm

AND endovascular surgery

Supplementary table (1): demographics of included studies with mid-term and long-term outcomes of elective fenestrated/branched endovascular aneurysm repair (F/BEVAR).

Abbreviations: Not reported (NR), Number of participants (n), maximum (max.), custom-made (CM) Thoracoabdominal aortic aneurysm (TAAA)

| **Author**  **/Year** | **Country and single- or multi-centre study** | **n** | **Mean follow-up (years) & period of data collection** | **Mean age (years)** | **Male (n)** | **Max. aortic aneurysm diameter (millimetres)** | **Juxtarenal aneurysms (n)** | **Pararenal aneurysms (n)** | **Suprarenal aneurysms (n)** | **Short-neck infrarenal aneurysms (n)** | **Type**  **I TAAA (n)** | **Type 2 TAAA**  **(n)** | **Type 3 TAAA**  **(n)** | **Type 4 TAAA**  **(n)** | **Type 5 TAAA (n)** | **Brand**  **of**  **fenestrated**  **/branched graft** | **CM or Off-the-shelf, Fenestrated (F), Branched (B)** | **N of target vessels per patient** |
| --- | --- | --- | --- | --- | --- | --- | --- | --- | --- | --- | --- | --- | --- | --- | --- | --- | --- | --- |
| Barilla 2014 | France, single | 50 | 5, 2006-2010 | 71.8 | 48 | NR | -- | -- | -- | 50 | -- | -- | -- | -- | -- | NR | NR, F | 2 |
| Kristmundsson 2014 | Sweden, single | 54 | 6.6, 2002-2007 | 72 | 46 | 60 | 54 | -- | -- | -- | -- | -- | -- | -- | -- | Zenith device (William Cook Europe) | NR, F | 2.5 |
| Nessvi 2014 | Sweden, single | 81 | 3.3, 2007-2011 | 72 | 63 | 55 | 81 | -- | -- | -- | -- | -- | -- | -- | -- | Zenith stent graft (Cook Europe) | NR, F | 2.1 |
| Mastracci 2015 | USA, single | 610 | 8, 2001-2013 | 75.2 | 501 | NR | 258 | -- | -- | -- | --- | -- | -- | 352 | -- | NR | NR, F | NR |
| Motta 2019 | USA, single | 223 | 3, 2012-2017 | 72 | 72 | 60 | -- | -- | -- | -- | Specific types of thoracoabdominal aneurysms not specified. | | | | | Zenith fenestrated device (ZFEN, Cook Medical) | NR, F, B | 3.1 |
| Walker 2019 | USA, single | 146 | 3, 2006-2016 | 72.6 | 107 | 67 | -- | 33 | -- | -- | 6 | 31 | 18 | 42 | 12 | Zenith stent graft (Cook Medical) | CM, B | 3.7 |
| Arnaoutakis 2020 | USA, single | 92 | 3, 2002-2018 | 72 | 57 | 66 | -- | -- | -- | -- | -- | 24 | 48 | -- | -- | Zenith stent graft (Cook Medical) | NR, F, B | 2.7 |
| Dossabhoy 2020 | USA, single | 213 | 6.2, 2010-2018 | 74 | 147 | 64 | 90 | 27 | -- | -- | -- | -- | -- | 96 | -- | Zenith stent graft (Cook Medical) | NR, F, B | 2.8 |
| Gargiulo 2020 | Italy, single | 98 | 3, 2008-2019 | 73 | 95 | 58 | 98 | -- | -- | -- | -- | -- | -- | -- | -- | Zenith stent graft | NR, F | 3.7 |
| Pini 2020 | Italy, single | 243 | 3, 2010-2019 | 73 | 209 | 61 | 160 | -- | -- | -- | 83 | -- | -- | -- | -- | Cook Medical | NR, F, B | NR |
| Diamond 2021 | USA, single | 299 | 3, 2010-2018 | 74 | 208 | 64 | 114 | 37 | -- | -- | 1 | 32 | 54 | 57 | 4 | Zenith stent graft | CM, F, B | 3.2 |
| Gallitto 2021 | Italy, single | 147 | 3.1, 2006-2018 | 73 | 133 | 60 | 72 | 46 | -- | -- | -- | -- | -- | 29 | -- | Zenith stent graft | CM, F | 3.6 |
| Sveinsson 2021 | Denmark, single | 94 | 7.4, 2007-2011 | 71 | 71 | 44 | 94 | -- | -- | -- | -- | -- | -- | -- | -- | Zenith stent graft | NR, F | 3 |
| Forbes 2023 | Canada, single | 242 | 3.3, 2007-2020 | 74.9 | 185 | 65.6 | 161 | -- | -- | -- | 3 | 17 | 23 | 33 | 5 | Cook Medical, Vascutek (Terumo aortic) | CM, off-the-shelf, F, B | 3 |
| Aucoin 2023 | USA, Netherlands, Multi-centre | 1681 | 3, 2005-2020 | 73.7 | 1193 | 64.3 | 282 | -- | -- | 220 | 51 | 320 | 251 | 533 | 22 | Cook Medical | CM, F, B | NR |
| Tinelli 2023 | Italy, France,Multi-centre | 102 | 5.6, 2010-2016 | 71.8 | 97 | 59.8 | 36 | 63 | -- | -- | -- | -- | -- | 3 | -- | Cook Medical | CM, F | 2.5 |
| Katsargyris 2023 | Germany, single centre | 349 | 4.1, 2010-2020 | 72.3 | 313 | NR | 240 | -- | 62 | 47 | -- | -- | -- | -- | -- | Cook Zenith | CM, F, B | NR |
| Prendes 2024 | USA, Sweden, Germany, multi-centre | 729 | 3.4,2007-2022 | 74.5 | 560 | 59 | 316 | -- | 171 | 60 | -- | -- | -- | 181 | -- | Not specified | CM, F | 3.1 |
| Mesnard 2024 | USA, Netherlands, multi-centre | 463 | 3.2, 2013-2023 | 74 | 327 | NR | -- | 135 | -- | -- | 24 | 112 | 48 | -- | -- | Cook Medical | CM, off-the-shelf, F, B | 3.9 |
| Vallabhaneni 2024 | UK, multi-centre | 403 | 3.5, 2017-2019 | 75.8 | 359 | NR | 403 | -- | -- | -- | -- | -- | -- | -- | -- | Zenith fenestrated, fenestrated Anaconda, Jotec Extra design service) | NR, F | NR |
| Oderich 2024 | USA, multi-centre | 1109 | 3, 2005-2020 | 73.3 | 741 | 62 | -- | -- | -- | -- | 589 (types I-III) | | | 520 | -- | Cook Medical | CM, F, B | 4 |
| Kanamori 2025 | USA, single | 342 | 5.25, 2013-2019 | 74.8 | 241 | 63 | 119 | -- | -- | -- | 14 | 79 | 35 | 93 | -- | Zenith stent graft (Cook Medical) | CM, F, B | 3.7 |
| Yadavalli 2025 | VISION registry, multi-centre | 891 | 5, 2014-2019 | 75.1 | 663 | 58 | 755 | -- | -- | -- | -- | -- | -- | 136 | -- | Zenith stent graft (Cook Medical) | CM, off-the-shelf, F | 3.1 |
| Rossillon 2025 | France, single | 169 | 3.6, 2007-2021 | 74.5 | 157 | 59 | 169 | -- | -- | -- | -- | -- | -- | -- | -- | Zenith stent graft (William A. Cook) | CM, F | 3.8 |

Supplementary table (2): co-morbidities, intra-operative characteristics, post-operative complications and definition of reintervention of included studies with mid-term and long-term outcomes of elective fenestrated/branched endovascular aneurysm repair (F/BEVAR).

Abbreviations: Diabetes Mellitus (DM), Not reported (NR), Number of participants (n), Hypertension (HTN), Ischaemic heart disease (IHD), Heart failure (HF), Chronic kidney disease – Stage 3 or above (Stage 3 or above CKD), Chronic obstructive pulmonary disease (COPD), Hypercholesterolaemia (HC), Cerebrovascular disease (CVD), Number of participants (n), Gastrointestinal (GI), Superior mesenteric artery (SMA)

|  | **Co-morbidities** | | | | | | | | | **Intra-operative details** | | |  | **Post-operative complications (<30 days)** | | | |  |
| --- | --- | --- | --- | --- | --- | --- | --- | --- | --- | --- | --- | --- | --- | --- | --- | --- | --- | --- |
| **Author**  **/Year** | **DM**  **(n)** | **HTN**  **(n)** | **IHD(n)** | **HF**  **(n)** | **Stage 3 or**  **above CKD(n)** | **COPD**  **(n)** | **Smoker**  **(n)** | **HC**  **(n)** | **CVD**  **(n)** | **Procedure time**  **(minutes)** | **Fluoroscopy time**  **(minutes)** | **Contrast volume**  **(ml)** | **Hospital-stay**  **(days)** | **Cardiac**  **(n)** | **Respiratory (n)** | **GI (n)** | **Renal (n)** | **Definition of reintervention** |
| Barilla 2014 | 12 | -- | 29 | -- | 8 | 24 | 36 | 25 | 3 | -- | -- | 138 | 12.0 | 1 | 1 | -- | 1 | Target vessel stenosis,haematoma, non-vascular interventions |
| Kristmundsson 2014 | 7 | 34 | 19 | 7 | 24 | 21 | NR | NR | NR | 250 | 78 | 270 | NR | NR | NR | 1 | NR | Interventions for endoleaks, target vessel stenosis/occlusion, graft limb stenosis/occlusion, extensions, thrombolysis/thrombectomy |
| Nessvi 2014 | 3 | 76 | 46 | NR | NR | NR | 38 | NR | 14 | NR | NR | NR | NR | NR | NR | NR | NR | Not specified |
| Mastracci 2015 | 119 | NR | 333 | NR | NR | 190 | 113 | NR | NR | NR | NR | NR | NR | NR | NR | NR | NR | Device body, endoleaks (number of reinterventions only) |
| Motta 2019 | 43 | 206 | 118 | 38 | 93 | 132 | 211 | 178 | 33 | 264 | NR | NR | 4.7 | 3 | 12 | 7 | 11 | Endoleaks, stent extensions, angioplasty |
| Walker 2019 | 17 | NR | 75 | NR | 46 | 78 | 129 | NR | 37 | NR | NR | NR | NR | NR | NR | NR | NR | Endoleaks, target vessel stenosis/occlusion, graft limb occlusion, graft explant |
| Arnaoutakis 2020 | 17 | 90 | 39 | NR | 35 | 53 | 76 | 77 | 5 | 220 | NR | NR | NR | 4 | 5 | 2 | 5 | Not specified |
| Dossabhoy 2020 | 31 | 195 | 102 | NR | 108 | 79 | 62 | 182 | 28 | 430 | 67 | 66 | 3 | 1 | NR | NR | 41 | Not specified |
| Gargiulo 2020 | 17 | 87 | 37 | NR | 33 | 39 | NR | 70 | 13 | 330 | 67 | 115 | NR | 6 | 8 | 22 | NR | Target vessel stenosis/occlusion, endoleak |
| Pini 2020 | 29 | 226 | 88 | NR | 100 | 105 | 82 | 226 | NR | NR | NR | NR | NR | 4 | NR | NR | NR | Not specified |
| Diamond 2021 | 46 | 278 | 143 | -- | 24 | 108 | 230 | 258 | 38 | 240 | NR | 69 | 4 | 22 | 1 | NR | 19 | Endoleaks |
| Gallitto 2021 | 25 | 130 | 56 | -- | 55 | 58 | NR | 107 | 19 | NR | NR | NR | NR | 5 | 6 | 1 | 24 | Endoleaks, target vessel stenosis/occlusion |
| Sveinsson 2021 | 10 | 61 | 36 | NR | 9 | 28 | 66 | NR | 9 | NR | NR | NR | NR | NR | NR | NR | 1 | Endoleak, target vessel stenosis/occlusion, graft explant, laparotomies, others |
| Forbes 2023 | 44 | 206 | 110 | 24 | 100 | 99 | 203 | 181 | 40 | 435 | 111 | 180 | 6 | 13 | 17 | 11 | 27 | Not specified |
| Aucoin 2023 | 230 | 1529 | 765 | 208 | 25 | 566 | 1464 | NR | 197 | 250 | NR | 107 | NR | NR | NR | NR | 120 | Not specified |
| Tinelli 2023 | 13 | NR | 43 | NR | 25 | 41 | 30 | NR | NR | 160.9 | 81 | 148 | NR | 4 | 6 | NR | 30 | Endoleaks, target vessel stenosis, graft explant |
| Katsargyris 2023 | 61 | 287 | 225 | NR | NR | 143 | 180 | 154 | NR | NR | NR | NR | NR | NR | NR | NR | NR | Endoleak, graft explant, target vessel stenosis/occlusion, graft limb extensions |
| Prendes 2024 | 92 | 638 | 258 | NR | 95 | 198 | 230 | 258 | 67 | 300 | NR | 198 | 7 | 26 | 23 | 16 | 107 | Endoleak, target vessel occlusion/stenosis, graft limb extensions |
| Mesnard 2024 | 65 | 414 | 222 | 41 | 202 | 138 | 360 | 365 | 46 | 230 | 79.2 | 152 | 6.7 | 13 | 17 | 11 | 27 | Endoleaks (in text), no numbers |
| Vallabhaneni 2024 | 78 | 338 | 196 | 46 | 75 | 159 | NR | NR | 19 | NR | NR | NR | 4 | 5 | NR | NR | 45 | Endoleak, target vessel stenosis/occlusion, conversion to open repair |
| Oderich 2024 | 143 | 1018 | 485 | 131 | 501 | 371 | 972 | NR | 128 | 390 | 78 | 102 | 5 | 26 | 40 | 13 | 111 | Figures for major and minor reinterventions provided |
| Kanamori 2025 | 55 | 29 | 241 | 56 | 67 | 110 | 295 | 309 | 37 | 241 | 81 | 148 | NR | 4 | 6 | NR | 30 | Endoleaks, target vessel occlusion/stenosis |
| Yadavalli 2025 | 178 | 786 | 198 | 157 | 53 | 326 | 811 | NR | NR | NR | NR | NR | NR | 30 | 40 | 1 | 127 | Not specified |
| Rossillon 2025 | 23 | 130 | NR | 15 | NR | 80 | 136 | 110 | NR | NR | 72.9 | 222 | NR | 2 | 4 | 9 | 27 | Endoleaks, lower limb, target vessel stenosis/occlusion |

Supplementary table (3): Robins-I (v2) risk of bias assessment of included studie^9^.

| Author/Year | Domain 1:  Risk of bias due to confounding | Domain 2: Risk of bias in classification of interventions | Domain 3: Risk of bias in selection of participants into the study | Domain 4: Risk of bias due to deviations from intended interventions | Domain 5: Risk of bias due to missing data | Domain 6: Risk of bias arising from measurement of the outcome | Domain 7: Risk of bias in selection of the reported result | Overall bias |
| --- | --- | --- | --- | --- | --- | --- | --- | --- |
| Barilla 2014 | low | low | low | low | moderate | low | low | low |
| Kristmundsson 2014 | low | low | low | low | low | low | low | low |
| Nessvi 2014 | low | low | moderate | low | moderate | low | moderate | low |
| Mastracci 2015 | low | low | low | low | low | low | low | low |
| Motta 2019 | low | low | moderate | low | moderate | low | moderate | low |
| Walker 2019 | low | low | low | low | moderate | moderate | moderate | moderate |
| Arnaoutakis 2020 | moderate | low | moderate | low | high | moderate | moderate | moderate |
| Dossabhoy 2020 | low | low | low | high | moderate | moderate | moderate | Moderate |
| Gargiulo 2020 | moderate | low | low | low | moderate | low | low | low |
| Pini 2020 | moderate | low | low | low | moderate | low | moderate | low |
| Diamond 2021 | moderate | low | low | moderate | moderate | low | low | low |
| Gallitto 2021 | moderate | low | low | low | moderate | moderate | moderate | moderate |
| Sveinsson 2021 | low | low | low | low | moderate | low | low | low |
| Forbes 2023 | moderate | low | low | low | moderate | moderate | moderate | moderate |
| Aucoin 2023 | moderate | low | moderate | low | high | moderate | moderate | moderate |
| Tinelli 2023 | low | low | low | low | low | low | low | low |
| Katsargyris 2023 | moderate | low | low | low | moderate | low | low | low |
| Prendes 2024 | moderate | low | moderate | low | moderate | low | moderate | moderate |
| Mesnard 2024 | low | low | low | low | moderate | moderate | low | low |
| Vallabhaneni 2024 | low | low | low | low | moderate | low | low | low |
| Oderich 2024 | low | low | low | low | low | moderate | moderate | low |
| Kanamori 2025 | low | low | low | low | moderate | low | low | low |
| Yadavalli 2025 | low | low | low | low | moderate | low | low | low |
| Rossillon 2025 | moderate | low | low | low | low | low | low | low |

Supplementary table (4). GRADE Analysis.

| **A contemporary systematic review and time-to-event analysis of mid- to long term outcomes in elective fenestrated/branched endovascular repair of complex abdominal and thoracoabdominal aortic aneurysms** | | | | |
| --- | --- | --- | --- | --- |
| **Patients or population:** Patients who underwent F/BEVAR for complex abdominal aortic and thoracoabdominal aneurysms  **Intervention:** F/BEVAR  **Follow-up:** up to 10 years  **Design:** all observational cohorts (single-arm), no randomised controlled trials | | | | |
| Outcomes | No. of participants (studies) | Effect estimate | Quality of the evidence (GRADE) | Comments |
| **All-cause mortality** | 24 studies, 8,886 patients | KM survival: 91.3% at 1 yr; 73.0% at 3 yrs; 55.4% at 5 yrs; 29.3% at 10 yrs. Median survival 6.36 yrs (Weibull). | ++OO  Low | (1) *Risk of bias:* all non-randomised, observational, varying follow-up, but no published randomised controlled trial in this area to date. (2) *Inconsistency:* two studies reporting mortality with cumulative incidence function out of 24 studies. (3) *Imprecision:* small numbers at risk beyond 5 years => wide CIs at 10 years. |
| **Freedom from aneurysm- related mortality** | 6 studies, 2,818 patients | KM survival: 92.4% at 1 yr; 84.5% at 3 yrs; 65.1% at 5 yrs. | ++OO  Low | (1) *Risk of bias:* observational (2) Outcomes reported from complex abdominal aneurysms, thoracoabdominal aneurysms and mixed cohort of aneurysm types. (3) Very small numbers at risk at 10 years. (4) Two studies reported mortality with cumulative incidence function (as above). |
| **Freedom from reintervention** | 15 studies, 4,555 patients | KM survival: 85.4% at 1 yr; 73.6% at 3 yrs; 66.5% at 5 yrs; 60.3% at 10 yrs. Median time to reintervention = 16.4 yrs (Logarithmic normal distribution). | ++OO  Low | (1) *Risk of bias:* observational reintervention definitions inconsistent; registry data with incomplete procedural coding. (2) *Imprecision:* late follow-up sparse; parametric model has wide CI. (3) *Inconsistency:* major variation in indications for reintervention across studies. |
| **Target vessel patency** | 3 studies, 1,714 patients | KM survival: 97.7% at 1 yr; 95.4% at 3 yrs; 94.9% at 5 yrs; 92.6% at 10 yrs. | + OO O  Very low | (1) *Risk of bias:* three single-centre studies with complete KM reporting only—may represent higher-performing centres. (2) *Imprecision:* small event numbers (patency >95% ⇒ very low events). |

Abbreviations: Confidence intervals (CI). Kaplan-Meier (KM). Log-likelihood ratio (LR). Year (yr). Years (yrs).

Supplementary Figure (1). Pooled versus individual Kaplan-Meier estimate of freedom from reintervention after F/BEVAR of complex abdominal aortic aneurysms.

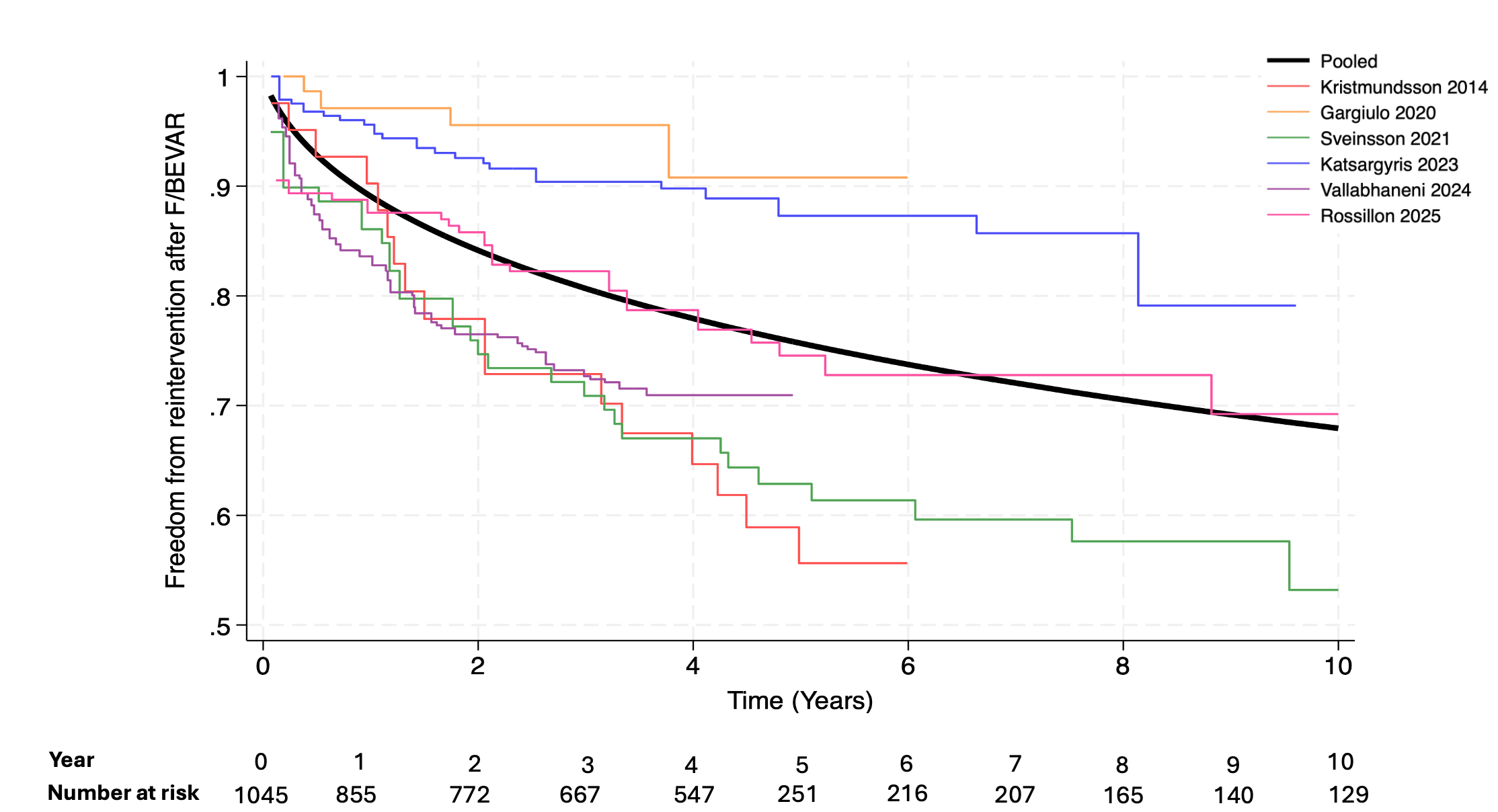

Supplementary Figure (2). Pooled versus individual Kaplan-Meier estimate of freedom from reintervention after F/BEVAR of thoracoabdominal aneurysms

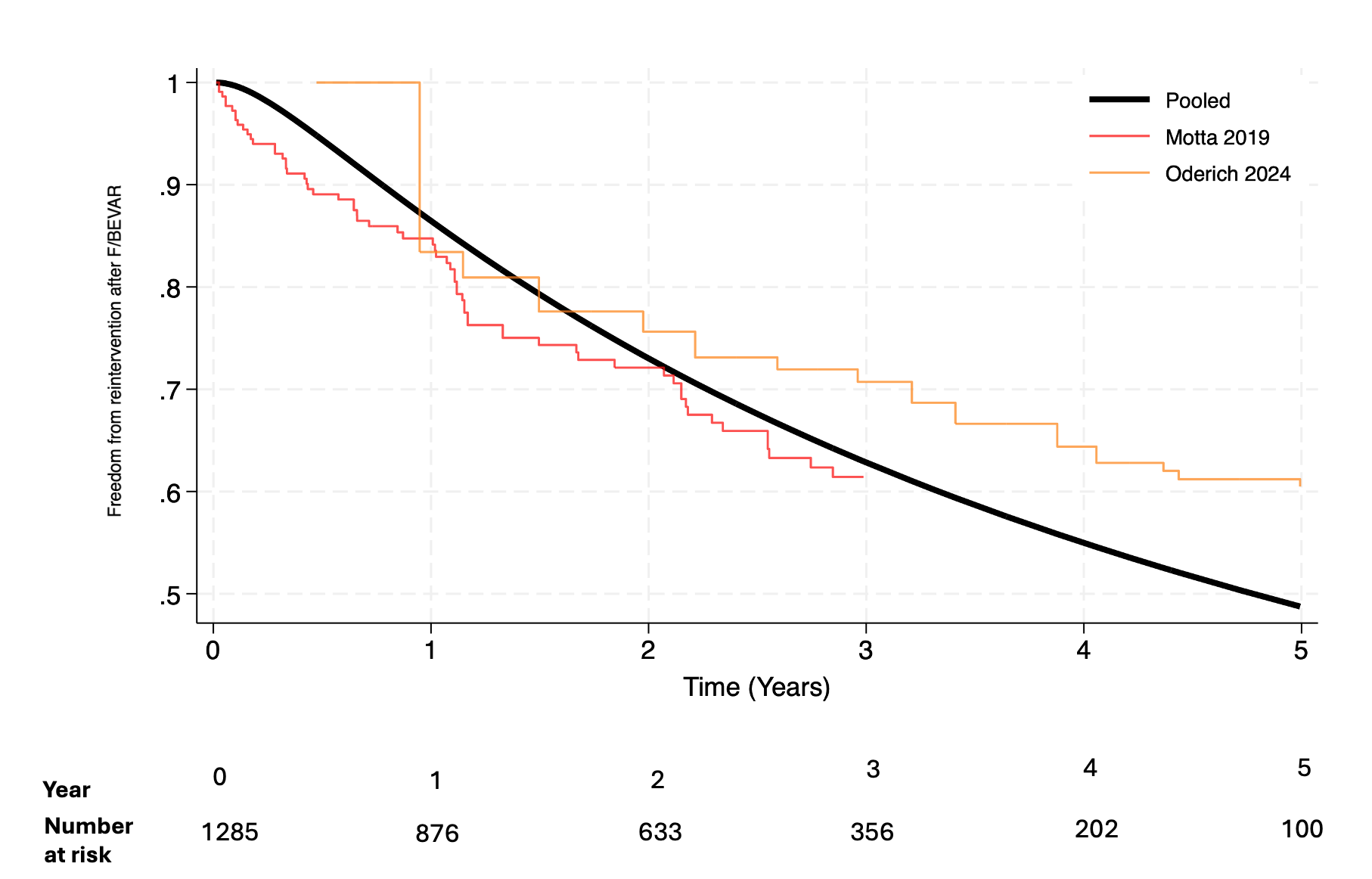

Supplementary Figure (3). Pooled versus Individual Target Vessel Patency after F/BEVAR

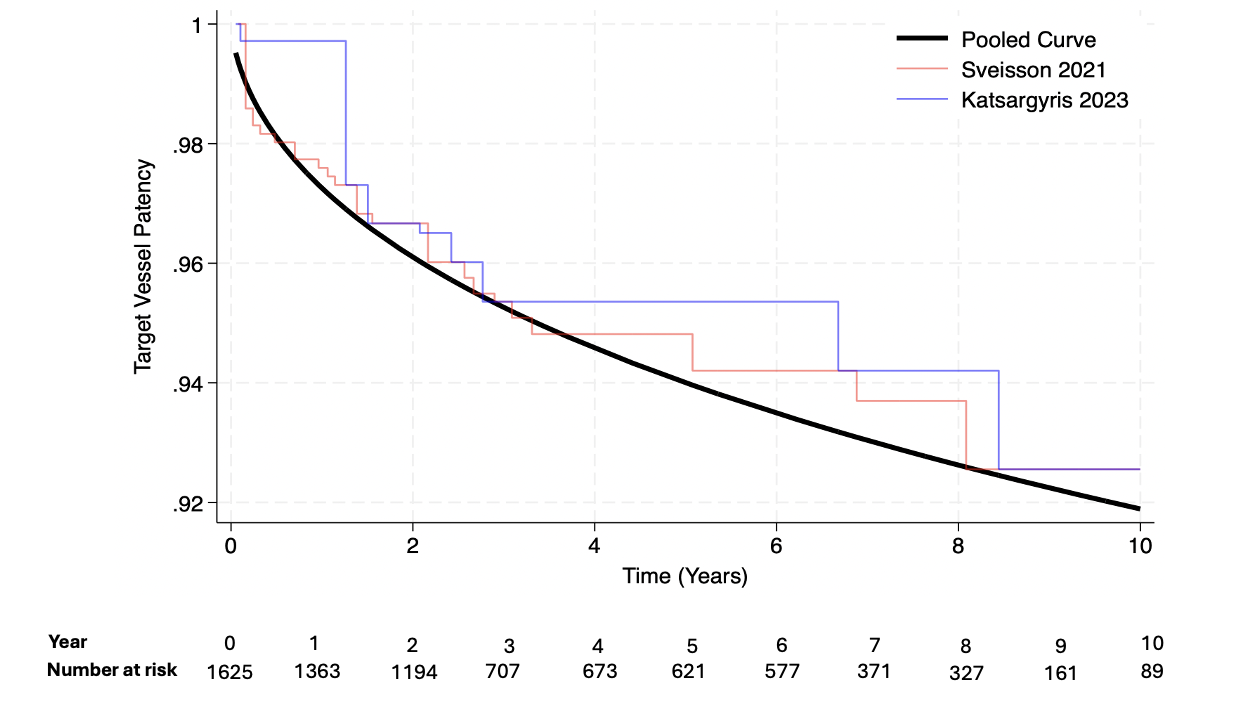
